# Text-message based assessment of 90-day modified Rankin Scale after Stroke

**DOI:** 10.1101/2023.11.14.23298546

**Authors:** Mohammad Hossein Abbasi, Kristy Yuan, Scott E. Kasner, Ellen McPartland, Karrima C. Owens, Kelly L. Sloane

**Author notes:** **Correspondence:** Kelly L. Sloane, Department of Neurology, 3W Gates Building, 3400 Spruce St., Philadelphia, PA 19104-4283.

## Abstract

**Background:** The modified Rankin scale (mRS) is commonly used to measure disability after stroke, traditionally assessed through telephone or in-person evaluation. Here, we investigated the validity of mRS assessment through automated text-messaging as an alternative method to traditional assessments.

**Methods:** Two hundred and fifty patients admitted to 3 hospitals within the University of Pennsylvania Health System with ischemic or hemorrhagic stroke were enrolled. Participants received automated text-messages sent 48 hours prior to their outpatient appointment at about 90-days post-stroke. The mRS scores were assigned based on participant responses to 2-4 text questions eliciting yes/no responses. The mRS was then evaluated in-person or by telephone interview for comparison. Responses were compared with kappa(κ).

**Results:** One hundred and forty-two patients (57%) completed the study. Spontaneous response rate to text messages was 46% and up to 72% with an additional direct in-person or phone call reminder. Agreement was substantial (quadratic-weighted κ=0.87) between responses derived from the automated text messaging and traditional interviews. Agreement for distinguishing functional independence (mRS 0-2) from dependence (mRS 3-5) was substantial (unweighted κ=0.79).

**Conclusion:** An automated text messaging system is a feasible and highly reliable for determining mRS and can serve as an alternative to traditional in-person or telephone assessment.

## Introduction

Stroke affects close to 800,000 individuals in United States annually and is the leading cause of adult disability in the United States ^1^. The modified Rankin scale (mRS) is a widely used outcome measure in clinical and research practices for assessing post-stroke disability ^2^. It is traditionally assessed through an in-person evaluation or structured telephone questionnaire ^3^, but both can be operationally difficult and time-intensive, and appointments or calls may be missed. Text-messaging may represent a user-friendly, accessible method for obtaining patient-reported outcomes. Prior text-message interventions for patient follow-up have been implemented for a variety of indications related to clinical follow-up and assessment of patient satisfaction including to surveillance of care for neurosurgical conditions ^4,5^, psychiatric disturbance ^6-8^, smoking and drinking cessation ^9,10^, diabetes mellitus management ^11,12^, and post-discharge primary care follow up ^13^. Text message-based approaches have grown in popularity over recent years, accelerated by the coronavirus pandemic and the essential need for strategies of remote patient monitoring ^14-16^.

Previously, the simplified mRS questionnaire (smRSq) was validated for assessment of post-stroke disability status with good reproducibility noted among raters certified in evaluation of mRS ^17-19^ and those uncertified ^20^. We hypothesized that an automated text-message based on smRSq would be a valid method to remotely determine mRS score at 90 days post-stroke compared to conventional in-person or telephone interview. This study introduces an innovative method for remote assessment of post-discharge stroke outcomes.

## Methods

The data that support the findings of this study are available from the corresponding author upon reasonable request.

We designed a 90-day post-discharge intervention using automated texting for assessment of the mRS score in collaboration with the platform, WayToHealth (WTH). WTH is a web-based platform that supports technology-based interventions to support behavior change interventions. WTH is integrated with the electronic medical record supported by the Penn Center for Health Care Innovation and Center for Health Incentives and Behavioral Economics. With WTH, we customized an automated text-message algorithm to message patients at specified intervals after discharge. We enrolled patients at 3 hospitals of Penn Medicine in Philadelphia, PA between August 2022 and February 2023: Hospital of the University of Pennsylvania, Pennsylvania Hospital and Penn Presbyterian Medical Center. Patients were eligible if they were hospitalized with an ischemic or hemorrhagic stroke, they (or a surrogate) could receive and respond to text-messages in English. Participants were excluded if life expectancy <90 days. The Institutional Review Board at the University of Pennsylvania considered this quality improvement study exempt from review. Patients or surrogates were approached prior to discharge to confirm eligibility and provide verbal assent, and then opted in to receive text-messaging from WTH. Participants were able to decline participation in the program by responding “stop” at any time.

Participants received a text-message 48 hours prior to their scheduled appointment, ideally at 90 days but allowing 30-180 days after stroke. Based on smRSq questionnaire, WTH used branching logic to ask 2-4 yes/no questions and determine the mRS score^17^(Figure 1). If the participant did not respond within 24 hours of the initial message, a second automated text-message with the same content was sent. At the in-person visit, mRS was obtained through a scripted interview by an individual rater (M.H.A.) who was trained in obtaining the mRS score and was blinded to text-message responses. The interview consisted of questions regarding the patient’s ability to complete activities of daily living, mobility, and dependence. If the participant was unable to attend an in-person visit, a telephone assessment was performed using the same scripted interview and rater. In situations where the surrogate responded to messages on behalf of the patient, the same surrogate was interviewed for the in-person/telephone-based assessment.

**Figure 1:**
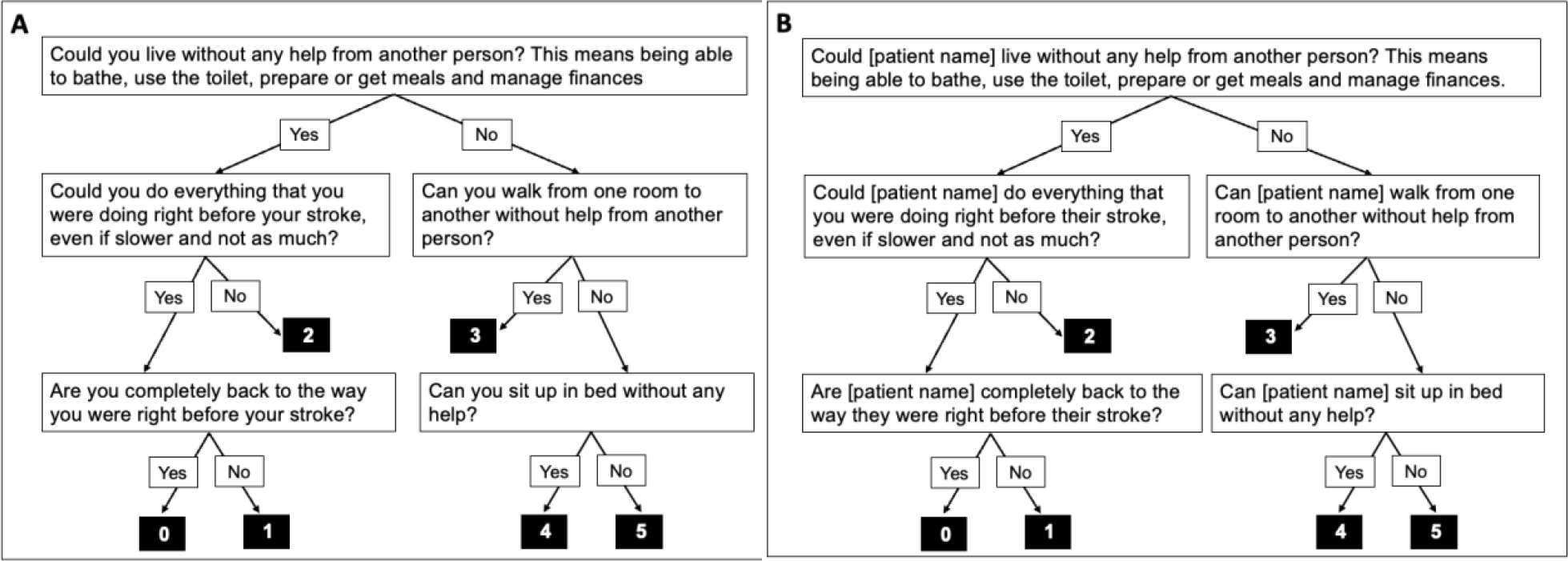
Automated text-message algorithm based on person providing response: modified based on Bruno et al. 2011. (Panel A: Patient as Responder Panel B: Surrogate providing Response).

Demographic and clinical characteristics were collected at baseline (Table 1). Participants also completed a survey of frequency of use and comfort with technology and living situation at 90-day follow-up.

**Table 1.** Demographic and clinical characteristics of cohort based on study completion.

|  |  | Enrollees (N=226) |  | P-value |
| --- | --- | --- | --- | --- |
|  |  | Completed study<br>(N: 142) | Did not complete<br>study<br>(N:84) |  |
| **Age, Median (Q1, Q3)** |  | 62 (54, 71) | 67 (59, 74) | 0.15 |
| **Female sex, n (%)** |  | 64 (45) | 35 (42) | 0.61 |
| **Education level, more than 12 years, n (%)** |  | 79 (56)* | NA | NA |
| **Patient as responder, n (%)** |  | 90 (63) | 58 (69) | 0.38 |
| **Race,**<br>**n(%)** | White | 68 (48) | 26 (31) | 0.11 |
|  | Black | 64 (45) | 46 (55) |  |
|  | Asian | 8 (6) | 2 (2) |  |
|  | Other races | 2 (1) | 2 (2) |  |
|  | Missing | 0 (0) | 8 (10) |  |
| **Ethnicity,**<br>**n(%)** | Hispanic or Latino | 7 (5) | 4 (5)* | 1.00 |
| **Stroke type,**<br>**n(%)** | Ischemic | 119 (84) | 77 (92) | 0.09 |
|  | Hemorrhagic | 23 (16) | 7 (8) |  |
| **Comorbidities,**<br>**n(%)** | DM with complications | 45 (32) | 40 (48) | 0.01 |
|  | Prior ischemic or<br>hemorrhagic stroke | 36 (25) | 21 (25) | 0.95 |
|  | CKD stage III or on dialysis | 10 (7) | 9 (11) | 0.33 |
|  | Complicated CAD or CHF<br>with NYHA classes III or IV | 24 (17) | 20 (24) | 0.20 |
|  | Neurodegenerative<br>conditions or spinal stenosis | 4 (3) | 1 (1) | 0.65 |
| **Pre-stroke mRS Median (Q1, Q3)** |  | 0 (0, 0) | NA | NA |
| **Baseline NIHSS Median (Q1, Q3)** |  | 4 (1, 8) | 4 (2, 7) | 0.86 |
| **90-day mRS (In-person) Median (Q1, Q3)** |  | 2 (1, 3) | NA | NA |
| NIHSS: National institute of health Stroke Scale, mRS: modified Rankin Scale, DM: Diabetes Mellitus,<br>CKD: Chronic Kidney Disease, CAD: Coronary Artery Disease, CHF: Congestive Heart Failure<br>*Missing education level for 9 participants and ethnicity data for 4 participants.<br>Student's t-test was used for continuous normally distributed variables, Wilcoxon ranked sum test for<br>ordinal variables, chi-squared tests for categorical variables. |  |  |  |  |

We assessed agreement between mRS scores derived from text-message and those from traditional methods (in-person or telephone) using Cohen’s kappa(κ), quadratic-weighted (κ_w_) for ordinal scores and unweighted for scores dichotomized as functionally independent (mRS 0–2) or dependent (mRS 3–5). Sensitivity and specificity of the dichotomized data were also calculated. Participant characteristics were compared between enrollees who completed the study and those who did not. P-value <0.05 was considered statistically significant. STATA version 5.0 (StataCorp LLC) was used for analyses.

## Results

Of 250 patients enrolled, 24 withdrew before receiving the study text-message. Complete mRS assessments were received from 105 (42%) based solely on the automated process and an additional 58 (23%) responded to the text-message mRS assessment after a brief reminder from a member of the study team. Baseline characteristics of those who completed the study and those who did not are described in Table 1. Those who responded via the fully automated process were younger, more likely to be white, had milder strokes, and had fewer chronic comorbidities than non-responders (Supplementary Table 1). Of these 163 whose mRS was assessed via text-messaging, 142 completed the traditional mRS interview and constituted the final study population (Figure 2). At study completion, 57% of participants reported electronic technology use without assistance or perceived difficulties but 17% perceived difficulties with the technology, 9% required assistance of another person, and 13% reported no technology use. The majority (85%) were living at home.

**Figure 2:**
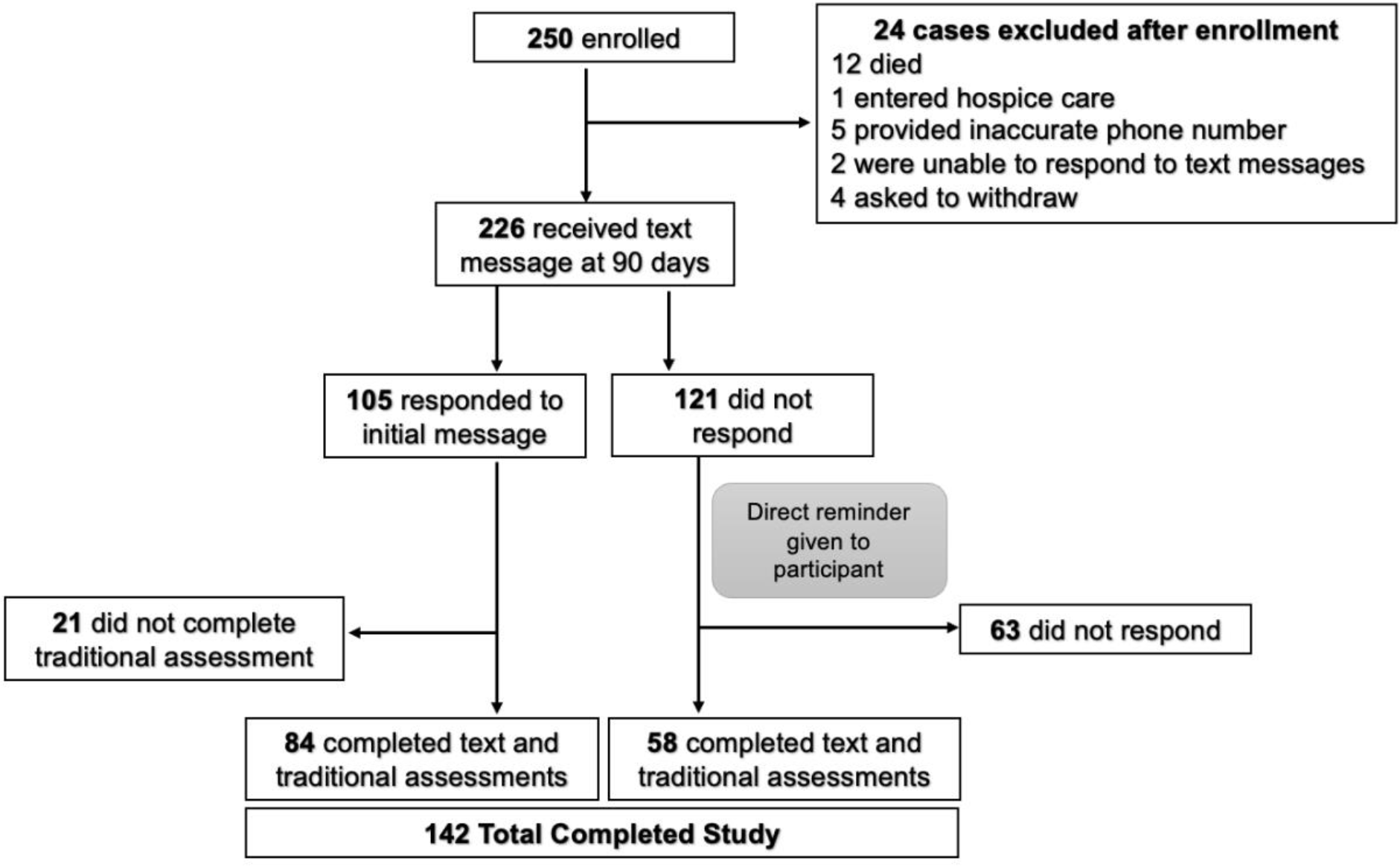
Study Flow Diagram

The final mRS measured through traditional methods (in-person or telephone) was a median of 2(1,3). There was excellent agreement between text-based and standard interviews (κ_w_=0.87, N=142) when evaluating ordinal mRS scores (Figure 3). Agreement with text-message-based mRS did not differ depending on method of assessment, in-person (κ_w_=0.86, n=69) or phone (κ_w_=0.87, n=73), nor by person responding, patient (κ_w_=0.86, n=90) or surrogate (κ_w_=0.78, n=52).

**Figure 3:**
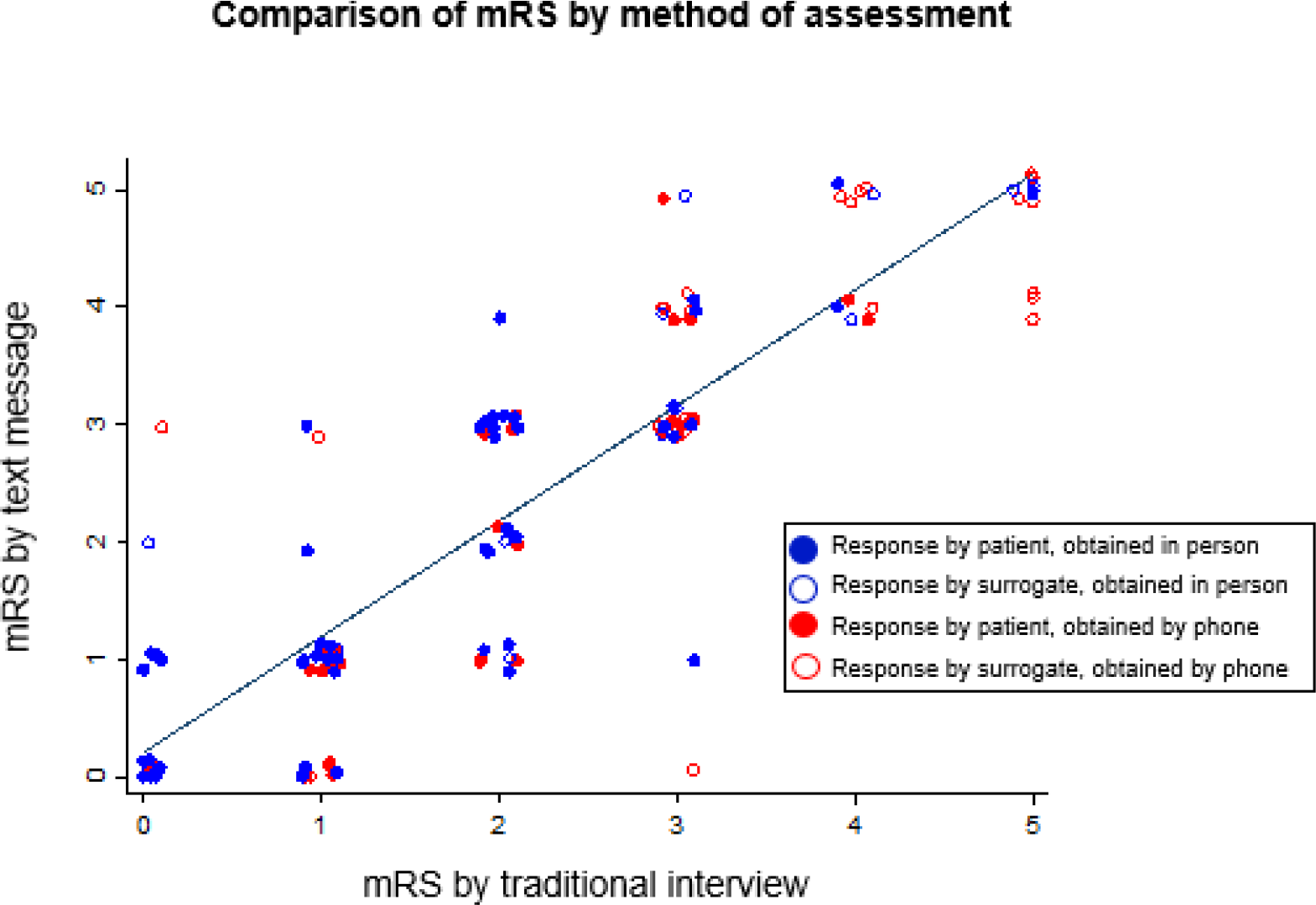
Comparison of modified Rankin Scale (mRS) by method of assessment (text message or traditional interview) and by person providing response (patient or surrogate).

With mRS dichotomized (mRS 0-2 vs. 3-5), there was substantial agreement between text-based and standard interviews. Discordant scores were assigned to 14 participants (Figure 3). Taking the standard interview as the reference criterion, the sensitivity of text-based assessment was 90% and specificity was 90%. Similar results were obtained for in person and phone assessments. There was greater agreement for dichotomized mRS when the patient responded compared to surrogates though confidence limits were overlapping.

## Discussion

Text-message communication is an emerging approach to patient engagement to support quality improvement and research. Text-messaging allows for automation and avoids common pitfalls of telephone or in-person assessments such as: unanswered calls, investment of time and effort among staff, and transportation and cost issues preventing patients from coming to clinic. As the mRS is commonly used to measure outcome after stroke in practice and in clinical trials, efforts to streamline and automate its collection should improve efficiency and access. Our study was launched as part of a quality improvement initiative to enhance assessment of outcome after recent stroke. We found that a text-message system of obtaining mRS at 90-days post-stroke was feasible, yielding a 42% spontaneous response rate. An additional 23% were willing to complete the process with an additional nudge. Text-based mRS had excellent agreement with standard interview-based mRS scores. Patient-reported mRS may have had better reliability between modalities than surrogate-reported scores. Further development may improve implementation and integration into clinical and research workflows. Automated text-messages from WTH can also be dispensed in multiple languages, and outcome data obtained from WTH can be incorporated into the electronic medical record.

Two recent studies have evaluated mRS through electronic or text communications and have found good reproducibility and validity. The first study described a smartphone/web application of the simplified mRS questionnaire, which would guide raters to a final mRS score. Sixteen raters from diverse clinical backgrounds participated, evaluating 24 different clinical vignettes. The study found strong agreement between the scores assigned by the individual raters and reference scores (weighted-κ=0.90) ^20^. Though this intervention is promising in terms of validity, it relies on the individual rater to assign an mRS score and therefore still requires the time and effort of a rater rather than an automated process.

Another more recent study assessed an automated text-message system similar to our program ^21^. They enrolled 350 participants to receive 3 sets of text messages at 30 days intervals post-discharge, out of which 85 patients completed the study up to the 90-day assessment. The intervention showed strong agreement between text-based and in-person assessments (weighted-κ=0.80) for both ordinal and dichotomized mRS scores. Although they had an initial number of 350 participants, they had a low response rate at 90-days of 22.8%. In addition to the low response rate, the study’s other limitations included the interval of 4 weeks between text-message response and in-person assessments and the lack of inclusion of surrogate participation. The post-stroke period reflects a dynamic time in recovery and there may be differences in function and level of disability that change rapidly between the first and second assessment. In addition, many individuals with recent stroke may not be able to receive and respond to text-messages, depending on their impairments and level of disability, so inclusion of surrogates will be an important aspect of validating an outcome measure.

In contrast to other published studies, our study involved narrow interval between types of assessments (in-person/telephone versus text-message based) with a higher response rate. We also sought to include surrogates as responders, as this reflects the real-world approach to mRS assessment where participants may not be able to self-report due to severity of disability. Our high spontaneous response rate was augmented by additional responses that occurred after a nudge from a human being, rather than a text-message, suggesting that some patients and caregivers may benefit from outreach that is not solely technology based. The platform used here, WayToHealth, can integrate with electronic medical records, allowing for the mRS to be auto-populated into the chart for review by the participant’s clinician.

Our study had several limitations. The participant sample was modest sized and was drawn from 3 hospitals all in the city of Philadelphia; therefore, generalizability to other populations may be limited. In addition, though our response rate was high among patients and their caregivers after discharge from stroke hospitalization, there were a substantial number of non-responders. Several patient-level factors may have contributed to this finding, including severity of illness, location of patient at 90 days (home or facility), comfort with technology and text-messaging, and access to a mobile phone. Rating of mRS for traditional assessments was performed by a single rater who obtained certification in this assessment, and there was no inter-rater reliability analysis given that mRS was assessed by a single rater.

Our study expands upon the evolving role of digital health applications in the care of patients with cerebrovascular disease. Text-message based outcome measures in this study yielded high response rates from patients and demonstrated substantial agreement with the traditional methods for obtaining these measures. Text-message based programs like WTH are a promising direction for post-discharge outcome assessments for stroke and other cardiovascular or cerebrovascular conditions. As a result of these promising findings, our institution is continuing to collect the mRS through tthis automated technology-based system. Future studies will be needed to determine which patient subgroups are less likely to respond to text-messaging and to determine if other outcomes measures may be validly collected through this platform.

## Acknowledgments

The authors would like to acknowledge Lisa Hernandes for her assistance in coordinating outpatient visits.

